# Effect of indoor air pollution on infant and child-mortality in Myanmar: Evidence from the first Demographic and Health Survey

**DOI:** 10.1101/19010801

**Authors:** Juwel Rana, Md Nuruzzaman Khan, Razia Aliani, Rakibul M Islam

## Abstract

**Background:** Indoor air pollution (IAP) from solid fuels for cooking has been considered as a public health threat, particularly for women and children in low- and lower-middle-income countries (LMICs). We investigated the effects of solid fuel use (SFU) on neonatal, infant and under-five child mortality in Myanmar.

**Materials and Methods:** We used data from Myanmar’s first Demographic and Health Survey conducted in 2016. The sample consists of ever-married mothers with under-five children in the household (n=3249). We calculated the adjusted odds ratio (aOR) to investigate the effects of SFU on neonatal, infant, and under-five mortality using multivariable logistic regression model accounting for survey weight and clustering. Additional analysis was conducted using an augmented measure of the exposure to IAP accounting for both SFU and the location of cooking (high exposure, moderate, and unexposed).

**Results:** The prevalence of SFU was 79.0%, and the neonatal, infant and the under-five mortality rates were 26, 45 and 49 per 1,000 live births, respectively. The odds of infant (aOR 2.17, 95% CI: 1.21, 3.88) and under-five child mortality (aOR 2.22, 95% CI: 1.24, 3.95) were higher in households with SFU compared with households with clean fuel use. When applying an augmented measure of exposure to IAP by incorporating both SFU and the kitchen’s location, the likelihood of infant and under-five mortality was higher among moderately and highly exposed children compared to unexposed children with similar trends. Neonatal mortality was not associated with both SFU and levels of exposure to IAP.

**Conclusion:** Infants and under-five children are at higher risk of mortality from exposure to IAP. The findings suggest that the risk of infants and under-five child mortality may be reduced by increasing access to clean cookstoves and clean fuels in LMICs, especially in Myanmar.

## 1. Background

Indoor air pollution (IAP) is one of the world’s major environmental threats, causing about 4 million premature deaths annually (1). IAP related mortality is disproportionately higher in low and middle-income countries (LMICs). In 2017, almost 70% of all deaths related to IAP occurred in LMICs (2). About 3 billion people use solid fuels for cooking, including coal and biomass (wood, animal dung, lignite, charcoal, wood, straw/shrubs, grass, and agricultural crop) (3,4), which are the major sources of IAP (1).

In contrast, alternative fuel (clean fuel) such as liquefied petroleum gas and electricity is often unavailable and/or unaffordable in LMIC (5). Therefore, households opt to collect solid fuels (3), which are burned indoors in conventional cookstoves as a pit, pieces of brick, or U-shaped mud construction. These solid fuels emit damaging airborne pollutants, including PM10, NOx, CO, SOx, formaldehyde and many toxic polycyclic aromatic hydrocarbons and other organic matter due to inefficient combustion (6–8). The amount of exposure to an individual in such settings has been measured to be much higher than the World Health Organization (WHO) guidelines and standards (9).

In LMICs, women and children are at higher risk of exposure to SFU (10,11) due to women’s role in household chores, cooking, and caring for infants in most South Asian culture. Women spend about three to seven hours per day near the stove, sometimes carrying their infants for care and warmth during cooking that leads children exposed to biomass fuel at similar levels (3). This exposure level increases in households with limited ventilation and poor design of the stove that do not have flues or hood to move out the smoke from living places (12).

The majority of the population in Myanmar use solid fuels for cooking as easy access to biomass for domestic cooking is a convention (13). Clean Cooking Alliance, Myanmar estimated that more than 95% of the rural and 88% of the urban population use solid fuels for cooking, which might be one of the causes of more than 3,500 yearly infant and children death from ALRIs and pneumonia in Myanmar (13,14). It could also be one of the reasons that Myanmar could not achieve the Millennium Development Goals (between 2000 and 2015) targets of reducing infant and child mortality (15). Importantly, this indicates an important area to address for achieving the Sustainable Development Goals of reducing neonatal (12 per 10000 live birth) and infant (25 per 10000 live births) deaths between 2015 and 2030.

No study, to our knowledge, has evaluated the effect of IAP from SFU on infant and under-five mortality rates using nationally representative data of Myanmar. The first Demographic Health Survey (DHS) in Myanmar was conducted in 2016 that provides an opportunity for us to examine the effect of SFU in the household on neonatal, infant and under-five mortality, comparing the magnitude of the effect based on the exposure levels.

## 2. Materials and Methods

### 2.1 Study design

Given the focus on improving maternal and child health, Myanmar Demographic and Health Survey (MDHS) 2016 was the first nationally representative survey conducted in Myanmar in 2015-16. Data were collected from 12,885 women from the sampled households based on stratified two-stage cluster sampling design. Using the 2014 Myanmar census sampling units, 442 clusters (123 urban, 319 rural) were selected in the first stage from 4,000 clusters through the probability proportional to the unit sizes. In the second stage, 30 households from each selected cluster were selected in the first stage by using systematic random sampling. The United States Agency for International Development (USAID) funded the DHS program, and the Ministry of Health and Sports, Myanmar, implemented MDHS 2016. Three Millennium Development Goals (a national level organization supported by the seven bilateral donors; Australia, Denmark, the European Union, Sweden, Switzerland, the United Kingdom and the United States of America). Technical support was provided by the ICF international. Detail of the survey sampling procedure has been published in the MDHS report (16).

### 2.2 Data Source

From 12,885 surveyed women, a sub-sample of 3,249 women were included in the analysis. The inclusion criteria were: i) women had at least one child born within five years before the date of survey; ii) respond to the question on survival status (alive/death at the time of survey) of this child; iii) respond to the question about the date of child death if applicable; and iii) respond to the question on household cooking fuels use.

### 2.3 Measures of Outcomes

We considered neonatal mortality (deaths occurred during the first 28 days of life), infant mortality (deaths occurred during the first one year (0-11 months) of life), and under-five mortality (deaths occurred during the first five years (0-59 months) of life) as outcome variables.

### 2.4 Measures of Exposure to IAP

The analysis was carried out for two exposure variables: Solid Fuel Use (type of clean fuel *vs*. solid fuel), and level of exposure to SFU induced IAP (unexposed, moderate exposure, and high exposure). The MDHS collected information on the type of cooking fuel by asking women, *what type of fuel does your household mainly use for cooking?* Responses were recoded as clean fuel =0 (if responses were electricity, liquid petroleum gas, and natural gas) and solid fuel =1 (if responses were coal, lignite, charcoal, wood, straw/shrubs, grass, and agricultural crop). Children’s levels of exposure to IAP was generated from the women’s responses to the place of cooking and the type of cooking fuel use. The responses were categorized as unexposed =0 (if women reported not using solid fuel), moderate exposure =1 (if women reported using solid fuel, but in a separate building or outdoors), and high exposure =2 (if women reported using solid fuel inside the house).

### 2.5 Confounder adjustment

Different sociodemographic factors that contribute to the neonatal, infant, and under-five mortality were also included as confounders. These were age at child deaths, child sex, women’s education, interval of last two succeeding births, household wealth quintiles, urbanity, geographic regions, and seasons. The birth interval variable was generated based on women’s response on the date of births of the last two-child and categorized by following the World Health Organization guidelines (16). The wealth quintile was generated from the women’s household durable and non-durable assets (e.g., televisions, bicycles, sources of drinking water, sanitation facilities and construction materials of houses) using principal components analysis.

### 2.6 Statistical analysis

Descriptive statistics were used to characterize the demographic profile of the respondents of the study. Differences in occurring neonatal, infant and under-five deaths across sociodemographic factors were also presented. The associations between exposure to IAP and child mortality outcomes were investigated using both univariable and multivariable logistic regression models. The univariable models included only the exposure variable and the outcome variable. These associations were then adjusted for potential confounders in the multivariable model. For each model, we used appropriate survey weights to adjust stratification and cluster variations in the survey data. Results are reported as odds ratios (OR) with 95% confidence interval (95% CI). All statistical tests were two-sided, and a *p*-value <0·05 was considered statistically significant. All analyses were carried out using statistical software packages Stata version 15.1 (Stata Corp: College Station, TX, USA).

## 3. Results

### 3.1 Sample characteristics

Characteristics of sample, exposures, and outcomes are presented in Table 1. The mean (SD) age of the mothers was 31(±6.0) years. The mean years of education was 4.43 (±3.5) years (SD±3.5). The mean age of the child was 2.11(±0.04) months, and nearly 48% of the child was female. More than three-quarter (77.8%) of the study households used solid fuel for cooking, of which 61.5% used solid fuel inside the house. About two-thirds (64.5%) of the women’s reported indoor place of cooking. Nearly half of the children and women (47.7%) were highly exposed to indoor air pollution during the survey (Table 1).

**Table 1:** Key information about the study participants, exposure and outcome variables

| **Demographics of mothers** | **Mean (SD)** |
| --- | --- |
| Mean age (±SD) | 31.09 (±6.0) |
| Mean weight (±SD) | 53.90 (±10.9) |
| Mean year of education (±SD) | 4.43 (±3.5) |
| **Demographics of children** | **Mean (SD)** |
| Mean age in months (±SD) | 2.11 (±0.04) |
| Female | 47.6 (45.4-49.8) |
| **Exposure to IAP** | **% (95% CI)** |
| Solid fuel use | 77.8 (74.1-81.1) |
| Solid fuel use (indoor only) | 61.5 (57.7-65.2) |
| Indoor cooking place (without SFU) | 64.5 (61.0-67.8) |
| **Level of exposure to IAP** | **% (95% CI)** |
| Unexposed | 22.4 (19.1-26.1) |
| Moderate exposure | 29.9 (26.8-33.0) |
| High exposure | 47.7 (43.9-51.6) |
| **Outcomes** | **(95% CI)** |
| Neonatal mortality per 1000 live birth | 26 (19-35) |
| Infant mortality per 1000 live birth | 45 (35-57) |
| Under-five mortality per 1000 live birth | 49 (38-62) |

SD= standard deviation, CI= confidence interval, IAP= indoor air pollution

The rate of neonatal mortality was 26 (95% CI: 19-53), infant mortality was 45 (95% CI: 35-57), and under-five mortality was 49 (95% CI: 38-62) per 1000 live births (Table 1). Infant and under-five child mortality were higher among children whose mother had no education, resided in Shan, Chin, and Teninthayi regions, and were born in short birth interval (Table 2).

**Table 2:** Child Mortality Outcomes by socio-demographic and spatial factors

| **Socio-demographic and spatial factors** | **Neonatal mortality** | **Infant Mortality** | **Under-five mortality** |
| --- | --- | --- | --- |
| **Age of the child, Mean(±SD)** | 0 (±0.24) | 1.74 (±0.26) | 3.93 (±0.65) |
| **Sex of child % (95% CI)** | **Neonatal mortality** | **Infant Mortality** | **Under-five mortality** |
| Male | 2.7 (1.9-3.9) | 4.4 (3.26.0) | 4.8 (3.5-6.5) |
| Female | 2.5 (1.6-4.0) | 4.6 (3.3-6.3) | 4.9 (3.6-6.7) |
| **Mothers' age at childbirth (Mean ±SD)** | 31.8 (±0.66) | 31. 48 (±0.53) | 31.78 (0.51) |
| **Mothers' education % (95% CI)** | **Neonatal mortality** | **Infant Mortality** | **Under-five mortality** |
| None | 4.4 (2.8-6.8) | 7.7 (5.2-11.2) | 8.3 (5.5-12.4) |
| Primary | 1.8 (1.1-3.1) | 3.6 (2.5-5.2) | 4.0 (2.9-5.6) |
| Secondary | 2.1 (1.2-3.7) | 2.9 (1.8-4.8) | 3.0 (1.9-4.9) |
| Higher | 5.1 (1.5-16.1) | 5.1 (1.5-16.1) | 5.1 (1.5-16.1) |
| **Wealth quintile % (95% CI)** | **Neonatal mortality** | **Infant Mortality** | **Under-five mortality** |
| Poorest | 1.9 (0.8-3.9) | 4.0 (2.4-6.6) | 4.2 (2.6-6.8) |
| Poor | 2.1 (1.1-4.1) | 3.2 (1.9-5.5) | 3.8 (2.2-6.6) |
| Middle | 3.2 (1.8-5.6) | 4.8 (3.1-7.3) | 5.4 (3.6-8.0) |
| Richer | 2.7 (1.6-4.8) | 5.5 (3.5-8.5) | 5.9 (3.9-9.0) |
| Richest | 2.8 (1.5-4.9) | 4.5 (2.9-7.1) | 4.5 (2.9-7.1) |
| **Urbanity % (95% CI)** | **Neonatal mortality** | **Infant Mortality** | **Under-five mortality** |
| Urban | 2.8 (1.6-4.8) | 4.3 (2.7-6.8) | 4.6 (2.9-7.1) |
| Rural | 2.6 (1.8-3.6) | 4.6 (3.4-6.1) | 5.0 (3.7-6.6) |
| **Geographic region % (95% CI)** | **Neonatal mortality** | **Infant Mortality** | **Under-five mortality** |
| Kachin | 2.4 (1.0-5.7) | 3.1 (1.4-7.0) | 3.5 (1.6-7.5) |
| Kayah | 2.2 (1.1-4.5) | 2.9 (1.5-5.6) | 2.9 (1.5-5.7) |
| Kayin | 1.9 (0.5-7.1) | 3.1 (1.2-7.6) | 3.5 (1.4-8.6) |
| Chin | 5.3 (3.6-7.9) | 7.5 (5.3-10.6) | 8.3 (5.5-12.2) |
| Sagaing | 2.4 (0.8-6.9) | 2.8 (1.1-7.2) | 3.2 (1.3-7.6) |
| Teninta | 1.7 (0.6-5.0) | 5.2 (2.0-12.7) | 6.9 (3.2-14.3) |
| Bago | 2.1 (0.8-5.7) | 3.3 (1.5-6.9) | 3.3 (1.5-6.9) |
| Magway | 2.4 (1.0-5.9) | 3.7 (1.9-6.7) | 4.3 (2.2-8.4) |
| Mandalay | 1.3 (0.3-5.0) | 3.8 (1.6-8.6) | 3.8 (1.6-8.6) |
| Mon | 1.8 (0.7-4.5) | 3.7 (1.6-8.1) | 4.3 (1.8-10.1) |
| Rakhine | 3.3 (1.4-7.6) | 3.8 (1.8-7.6) | 3.8 (1.8-7.6) |
| Yangon | 2.7 (0.6-11.9) | 4.3 (1.4-12.2) | 4.3 (1.4-12.2) |
| Shan | 3.8 (2.0-7.0) | 7.9 (4.5-13.5) | 8.4 (4.5-15.1) |
| Ayeyarwa | 3.2 (1.3-7.3) | 5.5 (2.8-10.3) | 6.0 (3.2-10.8) |
| Naypyiataw | 0.7 (0.09-4.0) | 2.0 (0.6-5.6) | 2.0 (0.7-5.7) |
| **Birth interval**<br>**% (95% CI)** | **Neonatal**<br>**mortality** | **Infant Mortality** | **Under-five**<br>**mortality** |
| ≥24 months | 2.31 (1.6-3.3) | 3.9 (3.0-5.0) | 4.3 (3.3-5.5) |
| <24 months | 4.7 (2.5-8.4) | 8.3 (5.1-13.1) | 8.8 (5.5-13.6) |
| **Seasons**<br>**% (95% CI)** | **Neonatal**<br>**mortality** | **Infant Mortality** | **Under-five**<br>**mortality** |
| Hot session | 1.5 (0.8-2.8) | 4.6 (2.8-7.5) | 5.1 (3.0-8.7) |
| Rainy session | 1.0 (0.3-4.0) | 1.8 (0.07-4.06) | 2.0 (0.83-4.7) |
| Cool session | 3.3 (2.3-4.6) | 4.8 (3.6-6.4) | 5.1 (3.9-6.7) |

### 3.2 Effect of exposure to IAP on neonatal, infant and under-five child mortality

The unadjusted and adjusted association between exposure and outcome variables are presented in Table 3. We found 2.22% (95% CI: 1.24-3.95) higher likelihood of under-five mortality, and 2.17 % (aOR 95% CI: 1.21-3.88) higher likelihood of infant mortality for the child of the women used solid fuel for cooking than women did not use solid fuel. The likelihoods were even higher when we considered the augmented measure of exposure to IAP. We found that the odds of occurring under-five mortality around 2.20 (95% CI: 1.19-4.06) and 2.25 (95% CI: 1.21-4.19) times higher among moderately and highly exposed children compared to unexposed. A similar higher likelihood of infant mortality was also observed among children with moderate (aOR 2.16 95% CI: 1.17-4.00) and high (aOR 2.19, 95% CI: 1.17-4.10) exposure IAP than their counterparts. There was no association between neonatal mortality with SFU and levels of exposure to IAP.

**Table 3:** Association between exposure to IAP and risk of neonatal, infant and under-five mortality in Myanmar

| Exposures | Mortality Outcomes |  |  |  |  |  |
| --- | --- | --- | --- | --- | --- | --- |
|  | Neonatal Mortality |  | Infant Mortality |  | Under-five Mortality |  |
|  | OR<br>(95% CI) | aOR<br>(95% CI)<sup>++</sup> | OR<br>(95% CI) | aOR<br>(95% CI)<sup>++</sup> | OR<br>(95% C)) | aOR<br>(95% CI)<sup>++</sup> |
| **Exposure to exposure** |  |  |  |  |  |  |
| **Clean fuel** | 1.00 | 1.00 | 1.00 | 1.00 | 1.00 | 1.00 |
| **SFU** | 1.55<br>(0.82-2.94) | 1.80<br>(0.77-4.21) | 1.68<br>(1.02-2.78)* | 2.17<br>(1.21-3.88)** | 1.85<br>(1.12-3.05) * | 2.22<br>(1.24-3.95)** |
| **Levels of exposure to IAP** |  |  |  |  |  |  |
| **Unexposed** | 1.00 | 1.00 | 1.00 | 1.00 | 1.00 | 1.00 |
| **Moderate** | 1.77<br>(0.88-3.58) | 1.98<br>(0.80-4.85) | 1.86<br>(1.07-3.23)* | 2.16<br>(1.17-4.00)* | 2.06<br>(1.19-3.56)** | 2.20<br>(1.19-4.06)** |
| **High** | 1.42<br>(0.72-2.79) | 1.57<br>(0.62-3.93) | 1.59<br>(0.94-2.70) | 2.19<br>(1.17-4.10)* | 1.74<br>(1.03-2.95)** | 2.25<br>(1.21-4.19)** |

\*p<0.05, \*\*p<0.01, aOR=adjusted odds ratios, CI= confidence interval; ++ Multivariable logistic models were adjusted for child age, gender, mothers' education, wealth quintiles, urbanity, geographic region, preceding birth interval and session.

## 4. Discussion

The risk factors of under-five child mortality, especially in the context of Myanmar, are under-investigated despite their high child mortality rate. Additionally, no study, to our knowledge, has examined the effect of IAP on child mortality in Myanmar. To address this gap, this study examined the associations of SFU and exposure to IAP with neonatal, infant, and under-five child mortality using first-ever nationally representative demographic and health survey data collected in 2016.

Our study suggests that the under-five mortality rate was high (48 deaths per 1,000 live births) in Myanmar. The majority of the households were dependent on SFU for cooking and heating purposes, and almost half of the study children were highly exposed to IAP in Myanmar. The study demonstrates that SFU and both moderate and high levels of exposure to IAP increased the risk of infant and under-five child mortality in Myanmar.

Previous studies reported comparable results that SFU and exposure to IAP increase the risk of infant and child mortality in LMICs (17–22). Evidence suggests that the combustion of SFU emits multiple pollutants such as fine particles, carbon monoxide, formaldehyde, and many more toxic chemicals, which increase the risk of mortality from ALRIs, asthma, and pneumonia among infant and young children exposed to these pollutants (3,6,7,23–29). Exposure to these toxic pollutants also increases the risk of low birth weight, stillbirth, and preterm birth, including acute and chronic health problems, all of which are considered as leading causes of child mortality (20,22,30,31).

Previous studies emphasized to consider cooking place along with SFU for examining the association between IAP and child-mortality because cooking inside the house with solid fuels maximizes the concentrations of airborne toxic pollutants in the household and ambient air (20,23,32,33). Following the previous study, we employed an inclusive SFU exposure measure combining both SFU and cooking place and found stronger associations of high exposure to IAP with infant and child mortality (23). Consistent with our study, previous studies showed that children were exposed to higher concentration of pollutants from SFU because of high proximity to pollutants and spending much time in the kitchen during heating and cooking, which intensifies the risk of child mortality from ALRI including other adverse health outcomes (17,23,24). The plausible explanation is that infants at their early age are often carried on their mother’s back or placed to sleep or stand beside their mother when they are cooking because this is common practice in South-East Asian countries, including Myanmar (20,21,32,34).

Furthermore, young children are more susceptible to IAP induced mortality compared to the older child for their underdeveloped epithelial linings of the lungs (23,35). However, women might be careful of IAP induced health effects on neonates. In a healthy condition, infants and young children have higher respiration, and they breathe 50% more polluted air due to their narrower airway and large lung surface. Also, children have a weak immune system in their early years of life which makes them vulnerable to IAP induced mortality, especially from ALRI (35–38).

However, neonatal mortality was not significantly associated with SFU and exposure to IAP, and these results are consistent with previous research conducted in LMICs (17,39). Several biological factors, such as low birth weight, prematurity, and complications associated with pregnancy and delivery, might be responsible for the null association between IAP and neonatal mortality (20,30,40). Additionally, breastfeeding could work as a protective factor diminishing the effect of IAP on neonatal mortality. Moreover, neonates and mothers might live in a conducive environment right after delivery, as well as mothers, usually stay away from any cooking activities during the neonatal period, which is also a cultural practice in Asia. However, few studies claim that neonates are at higher risk of IAP induced mortality and those were inconsistent across studies (22,41).

### Strength and weakness

The main strength of the study was based on the first-ever conducted a nationally representative survey with 98% response rate. Moreover, the analysis of large-scale data with appropriate statistical adjustments for confounders makes this study findings more reliable for policymaking. The temporal association between IAP exposure and child mortality outcomes cannot be established due to the cross-sectional nature of the study. Besides, the associations could be affected by unmeasured confounders as well as different health outcomes such as preterm birth, low birth weight, and other morbidity factors could be associated with under-five child mortality despite the exposure to IAP. Third, information related to birth and death of the children was reported by mothers that may introduce recall bias, but it is unlikely that the mother would incorrectly report the birth and death of their children. Fourth, there might be a source of exposure measurement error as we used two proxy measures such as SFU and combining SFU and cooking place to measure the associations between IAP exposures (23) and under-five child mortality. However, this is the available robust and established measurement of exposures for IAP because DHS does not objectively measure the level and duration of IAP exposures (23,32). Further studies may include questions related to ventilation in the kitchen, duration of cooking, proximity to the kitchen, or heating areas for the better measurement of children’s exposure to IAP.

## 5. Conclusion

The study demonstrates that SFU is a significant risk factor for infant and under-five child mortality but not for neonatal mortality. Furthermore, both moderate and high levels of exposure to IAP, such as the combination of SFU and cooking inside the kitchen, increase the risk of child mortality in Myanmar with a stronger effect among the children who had a high level of exposure to IAP. Policymakers should take both short-term and long-term strategies through socio-environmental pathways for addressing the problem of high child mortality rate in Myanmar. The government could decrease child mortality in Myanmar by implementing national policies related to clean fuels, cookstoves and green energy as well as reducing the level of exposures to IAP, which will ultimately help them to meet several sustainable development goals.

## Data Availability

DHS program gave us formal approval to obtain the de-identified data from the DHS online archive after. Details: https://dhsprogram.com/data/Using-325_DataSets-for-Analysis.cfm

https://dhsprogram.com/data

## Acknowledgments

The authors thank to MEASURE DHS for granting access to the first-ever conducted Myanmar Demographic and Health Survey (MDHS) 2016 data.

## Authors’ Contribution

JR and MNK developed the study concepts. JR and MNK analyzed the data. JR, MNK, and RA wrote the original draft of the manuscript. RI supervised and edited the manuscript. All authors critically read, reviewed and approved the final version.

